# Modifiable lifestyle factors and severe COVID-19 risk: Evidence from Mendelian randomization analysis

**DOI:** 10.1101/2020.10.19.20215525

**Authors:** Shuai Li

**Affiliations:** Centre for Epidemiology and Biostatistics, Melbourne School of Population and Global Health, The University of Melbourne, Parkville, Victoria, Australia; Centre for Cancer Genetic Epidemiology, Department of Public Health and Primary Care, University of Cambridge, Cambridge, United Kingdom

## Abstract

**Background:** Lifestyle factors including obesity and smoking are suggested to be related to increased risk of COVID-19 severe illness or related death. However, little is known about whether these relationships are causal, or the relationships between COVID-19 severe illness and other lifestyle factors, such as alcohol consumption and physical activity.

**Methods:** Genome-wide significant genetic variants associated with body mass index (BMI), lifetime smoking, alcohol consumption and physical activity identified by large-scale genome-wide association studies (GWAS) were selected as instrumental variables. GWAS summary statistics of these genetic variants for relevant lifestyle factors and severe illness of COVID-19 were obtained. Two-sample Mendelian randomization (MR) analyses were conducted.

**Results:** Both genetically predicted BMI and lifetime smoking were associated with about 2-fold increased risks of severe respiratory COVID-19 and COVID-19 hospitalization (all P<0.05). Genetically predicted physical activity was associated with about 5-fold (95% confidence interval [CI], 1.4, 20.3; P=0.02) decreased risk of severe respiratory COVID-19, but not with COVID-19 hospitalization, though the majority of the 95% CI did not include one. No evidence of association was found for genetically predicted alcohol consumption, but associations were found when using pleiotropy robust methods.

**Conclusion:** Evidence is found that BMI and smoking causally increase and physical activity causally decreases the risk of COVID-19 severe illness. This study highlights the importance of maintaining a healthy lifestyle in protecting from COVID-19 severe illness and its public health value in fighting against COVID-19 pandemic.

## Introduction

Obesity and smoking are lifestyle factors. Studies have reported the correlation between obesity and severe illness or related death of COVID-19^1-3^. For smoking, its relationship with the risk of severe COVID-19 is not clear^2,4,5^; however, these studies considered smoking as binary or categorical variables only without any consideration on the heaviness or duration. The Centre for Disease Control and Prevention of US suggests that people with obesity and smoking are at increased risk of COVID-19 severe illness^6^. The relationships are suggested by observational studies, which of themselves have limited ability in supporting causality. For other lifestyle factors, such as alcohol consumption and physical activity, little is known about if they are associated with severe illness of COVID-19.

Mendelian randomization (MR) uses exposure-associated genetic variants as instrumental variables to assess the causality between exposures and outcomes^7^. As genetic variants are randomly allocated at conception, MR resembles a randomized controlled trial and is less subject to confounding than observational studies. The publicly available genome-wide association studies (GWAS) summary statistics provide valuable resources for assessing the causality between lifestyle factors and the risk of COVID-19 severe illness. A MR study found evidence that both body mass index (BMI) and smoking had a causal effect on the risks of COVID-19 with respiratory failure and of hospitalization with COVID-19; however, the estimated causal effects had limited precision^8^.

This study aimed to investigate the causality between four lifestyle factors, namely BMI, smoking, alcohol consumption and physical activity, and severe illness of COVID-19 using the two-sample MR approach^9^.

## Methods

### COVID-19 data source

Summary-level data were obtained from two GWAS analyses conducted by the COVID-19 Host Genetic Initiative^10^ (Release 4 in September 2020): 1) 2972 very severe respiratory confirmed COVID cases, which were defined as hospitalized laboratory confirmed SARS-CoV-2 infection (RNA and/or serology based) with death or respiratory support, and hospitalization with COVID-19 as primary reason for admission, compared with 284472 population controls; and 2) 6492 hospitalized confirmed COVID cases, which were defined as hospitalized laboratory confirmed SARS-CoV-2 infection (RNA and/or serology based) and hospitalization due to corona-related symptoms, compared with 1012809 controls. The majority (≥90%) of the participants included in the GWAS analyses were of European ancestry. Details of the GWAS analyses can be found at https://www.covid19hg.org/

### Genetic instrumental variables to lifestyle risk factors

Genome-wide significant genetic variants identified from GWAS were selected as instrumental variables to the investigated lifestyle risk factors. BMI: 656 independent sentinel variants (P<10^−8^) identified from the meta-analysis of ∼700000 individuals of European ancestry from the Genetic Investigation of ANthropometric Traits (GIANT) consortium and UK Biobank, explaining ∼7.0% variation in BMI^11^. Smoking: 126 independent variants (P<5×10^−8^) identified from the GWAS of ∼460000 individuals from the UK Biobank for a lifetime smoking measure capturing smoking initiation, heaviness and duration, explaining 1.3% variation in the lifetime smoking measure^12^. Alcohol consumption: 81 independent sentinel (P<5×10^−8^) variants identified to be associated with drinks per week in a sample of ∼940000 individuals of European ancestry from the GWAS and Sequencing Consortium of Alcohol and Nicotine use (GSCAN), explaining ∼0.6% variation in alcohol consumption measured as drinks per week^13^. Physical activity: five variants identified to be associated with accelerometer-measured overall physical activity (measured as average vector magnitude) in a sample of ∼91000 UK Biobank individuals, explaining ∼0.2% variation in overall physical activity^14^. Proxies with a minimum linkage disequilibrium r^2^=0.8 were used for genetic variants that were unavailable in the COVID-19 data sources (two, one and two variants for BMI, alcohol consumption and physical activity, respectively; one alcohol consumption variant had no proxy available, so it was not included in analysis).

### Statistical analyses

The statistical power was calculated using the proportion of variation in the lifestyle risk factor explained by the genetic instrumental variables, sample size of the COVID-19 GWAS, and the method proposed by Burgess^15^.

The main analyses were performed using inverse-variance weighted (IVW) method under a random-effects model^16^, which assumes that all genetic variants are valid instrumental variables, or any horizontal pleiotropy must be balanced. For each risk factor, the reported odds ratio (OR) on COVID-19 risk was for per standard deviation increase in the genetically predicted value. Leave-one-out analyses, i.e., applying IVW after removing each genetic variant in turn, were performed to assess the influence of each genetic variant on the results.

Sensitivity analyses were performed using MR-Egger regression^17^, weighted median method^18^ and weighted mode method^19^, which relax some MR assumptions and allow some genetic instrumental variables to be invalid, but are less powerful than IVW method. The more consistency across the point estimates of the methods, the greater the evidence supporting the causal effect of the investigated risk factors on COVID-19 severe illness.

The analyses were conducted using the TwoSampleMR R package^20^. All statistical tests were two-sided. Results with a nominal P-value <0.05 were considered statistically significant.

## Results

For BMI, lifetime smoking, alcohol consumption and physical activity, respectively, this study has 80% statistical power at the significance level of 0.05 to detect an OR of 1.22, 1.57, 1.96 and 3.23 or greater on severe respiratory COVID-19, and an OR of 1.14, 1.36, 1.58 and 2.21 or greater on COVID-19 hospitalization.

Table 1 shows the causal effect estimates. Both genetically predicted BMI and lifetime smoking measure were found to be associated with increased risk of COVID-19 severe illness: the per-SD OR of genetic predicted BMI was 1.91 (95% confidence interval [CI], 1.55, 2.35, P=7.4×10^−10^) for severe respiratory COVID-19 and 1.75 (95% CI: 1.52, 2.01; P=9.0×10^−10^) for COVID-19 hospitalization; the per-SD OR of genetic predicted lifetime smoking was 1.84 (95% CI, 1.08 to 3.13, P=0.02) for severe respiratory COVID-19 and 2.15 (95% CI: 1.52, 3.03; P=1.5×10^−5^) for COVID-19 hospitalization. No evidence of association was found for genetic predicted alcohol consumption with severe respiratory COVID-19 or COVID-19 hospitalization. Genetically predicted physical activity was found to be associated with decreased risk of severe respiratory COVID-19 (OR=0.19; 95% CI: 0.05, 0.74; P=0.02), but not with COVID-19 hospitalization (OR=0.44; 95% CI: 0.18, 1.07; P=0.07), though most of the 95% CI did not included one. Similar results were found from the leave-one-out analyses, suggesting that the observed associations were not driven by any single genetic variant (Data not shown). There was evidence of heterogeneity in the genetic variant-exposure effects for BMI and alcohol consumption, but not for lifetime smoking and physical activity (Table 2).

**Table 1.** Odds ratios (OR) and 95% confidence intervals (CI) of the genetically predicted lifestyle factors with COVID-19 severe illness.

| Risk factor | Severe respiratory COVID-19 |  | COVID-19 hospitalization |  |
| --- | --- | --- | --- | --- |
|  | OR (95% CI) | P | OR (95% CI) | P |
| Body mass index | 1.91 (1.55, 2.35) | 7.4E-10 | 1.75 (1.52, 2.01) | 9.0E-15 |
| Lifetime smoking | 1.84 (1.08, 3.13) | 0.02 | 2.15 (1.52, 3.03) | 1.5E-05 |
| Alcohol consumption | 1.64 (0.70, 3.81) | 0.25 | 1.57 (0.90, 2.73) | 0.11 |
| Physical activity | 0.19 (0.05, 0.74) | 0.02 | 0.44 (0.18, 1.07) | 0.07 |

**Table 2.** Heterogeneity test results for the genetic variant-exposure effects.

| Risk factor | Method | Q | df | P |
| --- | --- | --- | --- | --- |
| Severe respiratory COVID-19 |  |  |  |  |
| Body mass index | Inverse variance weighted | 776.073 | 655 | 7.4E-04 |
| Body mass index | MR Egger | 770.065 | 654 | 1.1E-03 |
| Lifetime smoking | Inverse variance weighted | 137.387 | 125 | 0.21 |
| Lifetime smoking | MR Egger | 137.335 | 124 | 0.19 |
| Alcohol consumption | Inverse variance weighted | 118.041 | 79 | 2.9E-03 |
| Alcohol consumption | MR Egger | 110.051 | 78 | 0.01 |
| Physical activity | Inverse variance weighted | 3.273 | 4 | 0.51 |
| Physical activity | MR Egger | 3.005 | 3 | 0.39 |
| COVID-19 hospitalization |  |  |  |  |
| Body mass index | Inverse variance weighted | 772.906 | 655 | 9.7E-04 |
| Body mass index | MR Egger | 771.083 | 654 | 1.0E-03 |
| Lifetime smoking | Inverse variance weighted | 131.212 | 125 | 0.33 |
| Lifetime smoking | MR Egger | 131.192 | 124 | 0.31 |
| Alcohol consumption | Inverse variance weighted | 117.782 | 79 | 3.1E-03 |
| Alcohol consumption | MR Egger | 115.557 | 78 | 3.7E-03 |
| Physical activity | Inverse variance weighted | 3.629 | 4 | 0.46 |
| Physical activity | MR Egger | 1.852 | 3 | 0.60 |

From the sensitivity analyses (Figure 1), causal effect estimates with the same direction across the methods were found for all the investigated risk factors, though some estimates were with wider 95% CIs. Notably, positive associations (greater than the IVW estimates and the detectable effect sizes under 80% power) were found for alcohol consumption using methods other than the IVW method. Tests of MR-Egger regression intercepts suggested there was no evidence of directional pleiotropy of the genetic instrumental variables, with exception that weak evidence were found for BMI and alcohol consumption with severe respiratory COVID-19 (both P<0.03; Table 3).

**Table 3.** Test results for the MR-Egger regression intercepts.

| Risk factor | Intercept estimate | Intercept standard error | P |
| --- | --- | --- | --- |
| Severe respiratory COVID-19 |  |  |  |
| Body mass index | 0.010 | 0.004 | 0.02 |
| Lifetime smoking | 0.003 | 0.016 | 0.83 |
| Alcohol consumption | -0.024 | 0.010 | 0.02 |
| Physical activity | 0.108 | 0.210 | 0.64 |
| COVID-19 hospitalization |  |  |  |
| Body mass index | 0.004 | 0.003 | 0.21 |
| Lifetime smoking | 0.001 | 0.010 | 0.89 |
| Alcohol consumption | -0.008 | 0.007 | 0.22 |
| Physical activity | 0.162 | 0.096 | 0.19 |

**Figure 1.**
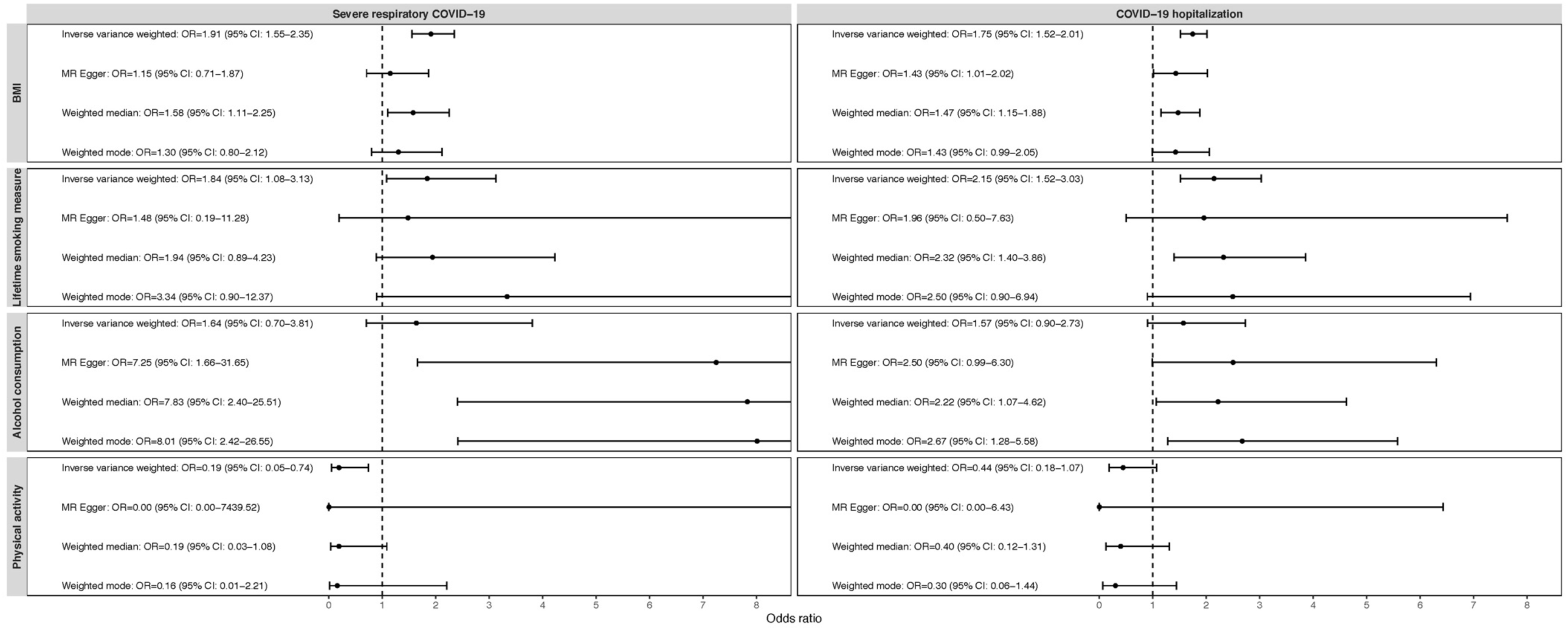
Odds ratios (OR) and 95% confidence intervals (CI) of the genetically predicted lifestyle factors with COVID-19 severe illness across methods. OR and 95% CI were expressed as per standard deviation increase in genetically predicted levels in body mass index (BMI), lifetime smoking measure, alcohol consumption (log-transformed standard drinks per week) and accelerometer-measured physical activity. The plots were right-truncated to better present the confidence intervals.

## Discussion

Using a two-sample MR approach, this study found evidence that BMI and smoking have a causal effect on increased risk of COVID-19 severe illness. These findings were the same as those by Ponsford et al.^8^, but the causal effect estimates were of greater precision than those by Ponsford et al., especially for smoking, e.g., the OR of smoking on COVID-19 hospitalization estimated by Ponsford et al. was 4.27 (95% CI: 2.10, 8.65), while this study estimated it to be 2.15 (95%: 1.52, 3.03). The difference is due to that this study used the summary statistics from COVID-19 GWAS analyses with larger sample sizes; Ponsford et al. used the summary statistics from a GWAS of severe COVID-19 with respiratory failure including 1610 cases and 2205 controls^21^, and the Release 3 data from the COVID-19 Host Genetic Initiative including 3199 cases and 897488 controls in the GWAS of COVID-19 hospitalization.

For the first time, this study provided evidence that physical activity causally decreases the risk of COVID-19 severe illness. However, only five genetic instrumental variables were used, and they explained ∼0.2% variation in physical activity only; the estimates for the causal effects were of reduced precision. The findings are perhaps more important in terms of qualifying causality than quantifying the causal effects.

As to alcohol consumption, this study had sufficient power to detect the observed IVW OR for COVID-19 hospitalization, but not for severe respiratory COVID-19. Interestingly, results from the MR-Egger regression, weighted median and weighted mode methods supported the association between genetically predicted alcohol consumption and COVID-19 severe illness. The consistency across the three methods suggest that the observed associations are unlikely to be biased by violated assumptions of a certain method. The three methods allow some genetic instrumental variables to be invalid. From the MR-Egger regression analyses, the intercepts were estimated to be negative, and the one for severe respiratory COVID-19 was even different from zero. Taking all these observations together, alcohol consumption might have a positive causal effect on COVID-19 severe illness, while the genetic instrumental variables overall might have negative directional pleiotropy, so the IVW results were not different from null.

Limitations of this study included that there might be bias in the causal effect estimates, as there was sample overlap between the lifestyle factors GWAS and COVID-19 GWAS, e.g., UK biobank participants were included in the GWAS of COVID-19 hospitalization. However, given the proportions of COVID-19 cases in the GWAS analyses were low, any bias must be minimal^22^.

In conclusion, this study finds evidence that BMI and smoking causally increase and physical activity causally decreases the risk of COVID-19 severe illness. All these lifestyle risk factors are modifiable, so they could be targeted to reduce severe illness of COVID-19. This study highlights the importance of maintaining a healthy lifestyle in protecting from COVID-19 severe illness. The findings also have a profound public health value – a healthy lifestyle could be helpful for fighting against the COVID-19 pandemic.

## Data Availability

The GWAS summary statistics used are publicly available.

## References

1. Docherty AB, Harrison EM, Green CA, et al. Features of 16,749 hospitalised UK patients with COVID-19 using the ISARIC WHO Clinical Characterisation Protocol. medRxiv 2020:2020.04.23.20076042.

2. Williamson EJ, Walker AJ, Bhaskaran K, et al. Factors associated with COVID-19-related death using OpenSAFELY. Nature 2020;584:430–6.

3. Simonnet A, Chetboun M, Poissy J, et al. High Prevalence of Obesity in Severe Acute Respiratory Syndrome Coronavirus-2 (SARS-CoV-2) Requiring Invasive Mechanical Ventilation. Obesity (Silver Spring, Md) 2020;28:1195–9.

4. Guan WJ, Ni ZY, Hu Y, et al. Clinical Characteristics of Coronavirus Disease 2019 in China. The New England journal of medicine 2020;382:1708–20.

5. Simons D, Shahab L, Brown J, Perski O. The association of smoking status with SARS-CoV-2 infection, hospitalisation and mortality from COVID-19: A living rapid evidence review with Bayesian meta-analyses (version 7). Addiction 2020.

6. Coronavirus Disease 2019 (COVID-19) - People with Certain Medical Conditions. 2020. (Accessed October 14, 2020, at https://www.cdc.gov/coronavirus/2019-ncov/need-extra-precautions/people-with-medical-conditions.html).

7. Smith GD, Ebrahim S. ‘Mendelian randomization’: can genetic epidemiology contribute to understanding environmental determinants of disease? International journal of epidemiology 2003;32:1–22.

8. Ponsford MJ, Gkatzionis A, Walker VM, et al. Cardiometabolic Traits, Sepsis and Severe COVID-19: A Mendelian Randomization Investigation. Circulation 2020.

9. Pierce BL, Burgess S. Efficient design for Mendelian randomization studies: subsample and 2-sample instrumental variable estimators. American journal of epidemiology 2013;178:1177–84.

10. The COVID-19 Host Genetics Initiative, a global initiative to elucidate the role of host genetic factors in susceptibility and severity of the SARS-CoV-2 virus pandemic. European journal of human genetics : EJHG 2020;28:715–8.

11. Yengo L, Sidorenko J, Kemper KE, et al. Meta-analysis of genome-wide association studies for height and body mass index in ∼700000 individuals of European ancestry. Human molecular genetics 2018;27:3641–9.

12. Wootton RE, Richmond RC, Stuijfzand BG, et al. Evidence for causal effects of lifetime smoking on risk for depression and schizophrenia: a Mendelian randomisation study. Psychological medicine 2019:1–9.

13. Liu M, Jiang Y, Wedow R, et al. Association studies of up to 1.2 million individuals yield new insights into the genetic etiology of tobacco and alcohol use. Nature genetics 2019;51:237–44.

14. Doherty A, Smith-Byrne K, Ferreira T, et al. GWAS identifies 14 loci for device-measured physical activity and sleep duration. Nature communications 2018;9:5257.

15. Burgess S. Sample size and power calculations in Mendelian randomization with a single instrumental variable and a binary outcome. International journal of epidemiology 2014;43:922–9.

16. Burgess S, Butterworth A, Thompson SG. Mendelian randomization analysis with multiple genetic variants using summarized data. Genetic epidemiology 2013;37:658–65.

17. Bowden J, Davey Smith G, Burgess S. Mendelian randomization with invalid instruments: effect estimation and bias detection through Egger regression. International journal of epidemiology 2015;44:512–25.

18. Bowden J, Davey Smith G, Haycock PC, Burgess S. Consistent Estimation in Mendelian Randomization with Some Invalid Instruments Using a Weighted Median Estimator. Genetic epidemiology 2016;40:304–14.

19. Hartwig FP, Davey Smith G, Bowden J. Robust inference in summary data Mendelian randomization via the zero modal pleiotropy assumption. International journal of epidemiology 2017;46:1985–98.

20. Hemani G, Zheng J, Elsworth B, et al. The MR-Base platform supports systematic causal inference across the human phenome. Elife 2018;7.

21. Ellinghaus D, Degenhardt F, Bujanda L, et al. Genomewide Association Study of Severe Covid-19 with Respiratory Failure. The New England journal of medicine 2020.

22. Burgess S, Davies NM, Thompson SG. Bias due to participant overlap in two-sample Mendelian randomization. Genetic epidemiology 2016;40:597–608.

